# Clinical malaria cases reflect the parasite population diversity of the asymptomatic reservoir in West Africa

**DOI:** 10.64898/2026.04.28.26351930

**Authors:** Bahdja Boudoua, Marc-Antoine Guery, Moussa Bamba Kanoute, Abdoulaye Katile, Mady Cissoko, Moussa Bamba Kanoute, Coralie L’Ollivier, Eva Legendre, Betty Kazanga, Jean Gaudart, Abdoulaye Djimde, El Hadj Konco Cire Ba, Issaka Sagara, Jordi Landier, Antoine Claessens

## Abstract

**Background:** Malaria transmission persists in Sahelian West Africa, with asymptomatic *Plasmodium falciparum* infections acting as a reservoir. Yet malaria epidemiology and research focus heavily on clinical cases. It remains unclear whether parasites causing clinical infections are genetically representative of the asymptomatic reservoir.

**Methods:** A one-year cohort study (2021–2022) was conducted in four villages in the Kedougou district of Senegal and two villages in the Kati district of Mali, collecting 1,133 *P. falciparum*-positive samples from asymptomatic and clinical infections. Using parasite molecular barcodes and whole-genome sequencing, we estimated multiplicity of infection (MOI), runs of homozygosity (RoH), and identity-by-descent (IBD) to compare parasite populations between infection types and across age groups.

**Results:** A total of 226 whole genomes and 728 barcodes were analysed, representing 78.2% (570/729) of clinical cases and 21.7% (158/728) of asymptomatic infections. MOI was high, with over 50% of infections being polyclonal, and did not differ by age or infection status. IBD analysis revealed a highly diverse parasite population, with transmission clusters including both asymptomatic carriers and clinical cases. Drug resistance allele frequencies were similar between groups.

**Conclusion:** Overall, we found no evidence of genetic differentiation between parasites from clinical and asymptomatic infections, suggesting that clinical cases reflect the broader parasite population sustaining transmission. These findings provide a basis for linking genomic metrics with epidemiological factors and for monitoring the impact of future interventions on the parasite population.

## Background

Despite decades of control efforts, malaria remains a significant public health and development challenge. In 2024, the WHO African Region reported 265 million cases and over 600,000 deaths, accounting for approximately 94% of global cases and 95% of global deaths. While substantial reductions of morbidity and mortality were achieved between 2000 and 2015, progress has since stalled in several moderate to high transmission settings despite continued deployment of control interventions (Doumbia et al. 2022; ‘World Malaria Report 2024’; Cissoko et al. 2025).

This global trend is reflected in the Sahelian region of West Africa, where malaria incidence decreased from 359 to 290 cases per 1,000 person-years between 2011 and 2018 before plateauing (Tairou et al. 2024; Sacko et al. 2021). Southeastern Senegal and southern Mali continue to experience high malaria prevalence despite comprehensive control programs (Cissé et al. 2016; Cissoko et al. 2022). Control strategies rely on coordinated awareness campaigns, universal coverage with long-lasting insecticidal nets (LLINs) distributed every 3 years, improving access to diagnosis and treatment, and seasonal malaria chemoprevention (SMC) campaigns targeting children aged 3-59 months in Mali and 3 months to 10 years in Senegal. In addition, one of the key interventions to reduce severe morbidity and mortality in Senegal and Mali was the deployment of SMC since 2012 (Manga et al. 2023).

In these regions, climate is characterised by a rainy season from late June to October, followed by an approximately 6-month dry season with minimal or no rainfall. During this dry period, when clinical cases and mosquito populations decline, asymptomatic carriers act as hidden reservoirs of infection, often harbouring gametocytes capable of infecting mosquitoes, thereby ensuring parasite persistence (Guery et al. 2025; Andolina et al. 2021). At the onset of the next wet season these reservoirs amplify transmission as mosquito populations rebound (Kazanga et al. 2026; Coulibaly et al. 2017). Despite their role in sustaining parasite circulation and their high prevalence, asymptomatic infections remain understudied and under detected. Malaria research has traditionally focused on clinical cases due to their immediate relevance to human health and the challenges in detecting low-density asymptomatic infections. Consequently, even though most *P. falciparum* infections are transmitted by asymptomatic carriers (Soumare et al. 2025), it remains unclear whether parasites causing clinical disease are representative of those in asymptomatic carriers (Searle et al. 2017). This uncertainty may compromise surveillance strategies that rely on either group alone and may lead to the mischaracterization of circulating parasite populations and the underestimation of asymptomatic transmission (Mwesigwa et al. 2024).

To address this gap, genomic epidemiology provides insights into parasite population structure and transmission dynamics that traditional surveillance methods cannot capture (Day et al. 2025). These approaches have been successfully used to detect associations between malaria parasite genetic diversity, dynamic changes in transmission intensity, and to assess impact of interventions (Sy et al. 2022).

This study aims to characterise the genetic epidemiology of *P. falciparum* circulating in both asymptomatic carriers and clinical malaria patients from the same communities in southern Senegal and southern Mali. Specifically, we calculate three complementary genomic metrics: Multiplicity of Infection (MOI) (Lopez and Koepfli 2021), Runs of Homozygosity (RoH), and Identity-by-Descent (IBD) (Guo et al. 2025), capturing within-host diversity and parasite relatedness at both the individual and population levels. We then compare them between infections identified as clinical cases or asymptomatic carriers, in order to assess whether clinical cases accurately represent the parasite reservoir, i.e. those in asymptomatic carriers, or whether asymptomatic carriers maintain distinct parasite lineages that contribute differently to transmission.

## Material and methods

### Study design and sample collection

We analysed samples from a previously reported cohort study in two sites: Kedougou Region of southeastern Senegal (4 villages located within a 20 km radius), and Kati Cercle, Koulikoro Region, southern Mali (the villages of Safo and Torodo, approximately 6 km apart and 15 km from Bamako) (Legendre et al. 2025; Kazanga et al. 2026). A third site in Northern Mali (Dire district, 2 villages) was excluded because no samples were collected from clinical malaria cases in that site (Kazanga et al. 2026).

In both Senegal and Mali, a household census was conducted in all participating villages, from which households were randomly selected. All consenting household members were eligible for cohort participation (Kazanga et al. 2026; Legendre et al. 2025). Cohort participants were surveyed during four scheduled visits spanning dry and wet seasons: T0 initial visit (April 2021 in Kedougou and May 2021 in Kati), T1 before seasonal malaria chemoprevention started (end June in Kedougou, end July in Kati), T2 peak prevalence (December 2021 in both sites), and T3 dry-season endline (April 2022 in Kedougou and May 2022 in Kati). At each visit, epidemiological data were collected and capillary blood samples were obtained by finger prick, with dried blood spots (DBS) prepared for laboratory analysis. Samples were extracted and tested for *P. falciparum* using varATS qPCR, as described previously (Kazanga et al. 2026; Legendre et al. 2025).

In addition to scheduled visits, cohort participants were asked to provide a DBS sample when diagnosed with RDT-confirmed clinical malaria at village health facility. In Senegal only, DBS collection was extended to all village inhabitants presenting with an RDT-confirmed malaria episode at health facilities. They are hereafter termed ‘non-cohort clinical cases’ and were analysed separately from cohort participants throughout this study. This additional sampling was not conducted in Mali.

### Ethics

Written informed consent was obtained from all participants, or from parents/legal guardians for those under 18 years old. In Senegal, inhabitants not included in the cohort provided specific informed consent for dried blood spot collection when they presented with a clinical malaria episode.

The study protocol received ethical approval from the National Ethics Committee for Health Research of Senegal (N°0000052/MSAS/DPRS/CNERS) and the Ethics Committee of the University of Sciences, Techniques and Technologies of Bamako, Mali (protocol No. 2020/297/CE/FMOS/FAPH).

### Definitions

#### Asymptomatic infection

An infection detected in individuals who do not actively seek treatment and do not present clinical symptoms but test positive for *P. falciparum* by varATS qPCR during the survey.

#### Clinical infection

An infection in individuals who actively present for diagnosis and treatment due to malaria-related symptoms and confirmed by RDT in health facilities.

#### Barcode

A short DNA sequence of 101 single polymorphic nucleotides (SNPs)

#### Genome

The complete set of DNA (genetic material)

#### Monoclonal infection

An infection in which a single parasite genotype is detected.

#### Polyclonal infection

An infection in which two or more distinct parasite genotypes are detected.

#### Co-transmission

Infection resulting from a single mosquito bite in which multiple genetically related parasite genotypes are transmitted simultaneously to an individual.

#### Superinfection

Infection resulting from bites by two or more mosquitoes, each transmitting a single parasite genotype, leading to the accumulation of distinct parasite genotypes within the same host.

#### Related pairs

parasite sample pairs sharing identity-by-descent (IBD) greater than 0.5 for barcode data and greater than 0.2 for whole-genome data.

#### Comparable sites

Pairs of homozygous sites between two samples

#### Informative sites

Comparable sites where at least one sample in the pair carries the minor allele, as defined at the population level.

### Plasmodium genomics and data analysis pipeline

*P. falciparum* positive DBS samples obtained either from RDT-confirmed clinical malaria patients or detected as *P. falciparum* positive by varATS qPCR were targeted for genotyping and whole-genome sequencing (WGS) at the Wellcome Sanger Institute with MalariaGEN. When the remaining amount of DBS was insufficient, extracted DNA was used directly. DBS extraction, selective whole-genome amplification, amplicon-based sequencing, and whole-genome sequencing were performed by the SpotMalaria platform (Oyola et al. 2016; da Silva et al. 2023) as described in (MalariaGEN et al. 2023). Details are provided in supplementary methods **(page 1, paragraphs 1-2)**. To analyse barcode data, together with WGS data formatted in a VCF format, we applied a previously developed pipeline (Guery et al. 2025), extended here with two additional metrics (effective MOI and Runs of Homozygosity):

#### WGS analysis pipeline

1. Filter out genomes that had <50% of bases covered at a minimum depth of 5 reads.
2. Filter out genomic loci with a population minor allele frequency below 0.1, with more than two alleles identified in the population, or located outside the core genome (Miles et al. 2016).
3. Estimate the proportion of polyclonal samples using the *F*ws metric and Runs of Homozygosity (RoH) (Pearson et al. 2016).
4. Format SNPs into a matrix as required by hmmIBD (Schaffner et al. 2018). The matrix was formatted as follows: 0 and 1 were used for the respective allele of each SNP and –1 for mixed and unknown positions (which were excluded from IBD estimation). SNP calls were considered mixed if the within-sample Minor Allele Frequency (MAF) exceeded 0.2 (Guery et al. 2025).
5. Use the paired IBD values obtained from hmmIBD and build a network of genome relatedness. IBD values were considered unknown between pairs of genomes having less than 100 informative sites.

#### Barcodes data analysis pipeline

1. Filter out barcodes that had ≥75 loci with no usable sequence data
2. Assessment of barcode genotyping quality The quality and accuracy of barcode genotyping were assessed by comparing molecular barcodes (constructed from genotyped single nucleotide polymorphisms, SNPs) with genomic barcodes (constructed from whole-genome sequencing) obtained from the same samples.
3. Estimate the proportion of polyclonal samples using the proportion of heterozygosity and the effective MOI (eMOI) (Murphy and Greenhouse 2024) (described in the following section).
4. Barcodes were formatted into a matrix as required by hmmIBD. Pairwise IBD estimates were calculated using hmmIBD, which applies a hidden Markov model to infer the proportion of the genome shared IBD between each pair of parasite samples (Schaffner et al. 2018).
5. Pairs of barcodes with insufficient genetic comparability were excluded, defined as those having fewer than 30 comparable sites (defined above) for monoclonal samples (eMOI ≤ 1.1) or fewer than 50 comparable sites for samples with 1.1 < eMOI ≤ 1.85, or fewer than 10 informative sites (defined above). For samples with an eMOI > 1.85, pairwise IBD values were treated as unknown and omitted from the network analysis.
6. The resulting filtered IBD matrix was then used to construct a network of parasite genetic relatedness.

### Estimating multiplicity of Infection

Multiplicity of Infection (MOI), also known as complexity of infection quantifies the number of distinct *P. falciparum* strains simultaneously infecting a single host.

We estimated MOI using multiple complementary approaches across different data types. For whole-genome samples, MOI was initially estimated using the Fws metric based on allelic frequencies (Auburn et al. 2012) and by quantifying long Runs of Homozygosity (RoH) (Pearson et al. 2016) as the primary metric, further described below as it informs the parasite relatedness analysis.

For barcode samples, we assessed MOI using two approaches: (i) the proportion of heterozygous loci (with infections classified as polyclonal if > 0.8% of sites were heterozygous, with a within-sample minor allele frequency cutoff of 0.2 (Guery et al. 2025)); and (ii) effective MOI (eMOI) as the primary metric used in downstream analyses, also described in below.

#### Effective MOI (eMOI)

For barcode samples, we relied on eMOI, which adjusts conventional MOI to account for genetic relatedness among parasite clones within an infection. This approach accommodates situations where parasites within a host are genetically related rather than independent. eMOI (Murphy and Greenhouse 2024) is defined as: eMOI=MOI×(1−r_w_)+1, where r_w_ represents the average genomic relatedness among clones within a host. Infections with eMOI > 1.1 were classified as polyclonal (multiple parasite genotypes), while those with eMOI ≤ 1.1 were considered monoclonal (single parasite genotype), a threshold supported by previous studies (Chen et al. 2025; Brokhattingen et al. 2024).

#### Runs of Homozygosity (RoH)

To obtain a more detailed picture of clone relationships within mixed infections from whole-genome data, we analysed long Runs of Homozygosity (RoH). A ‘run of homozygosity’, in an otherwise heterozygous infection, is a haplotype shared by all clones present within the infection. For an infection with two clones, a RoH across ∼50% of the genome indicates that the two clones are meiotic siblings. Lower percentages of RoH indicate a more distant relationship, while higher values indicate inbreeding over multiple generations.

RoH and genome-wide heterozygosity patterns were analysed using a custom Python script based on previously described approaches (Pearson et al. 2016; Fogang et al. 2025).

RoH were inferred from whole-genome sequencing data by first calculating the non-reference allele frequency (NRAF) at individual SNPs. SNP-level heterozygosity was then summarized in non-overlapping 20-kb windows across the genome. Contiguous windows showing consistently low heterozygosity were classified as RoH and visualized to identify extended homozygous segments.

NRAF patterns informed clone number, whereas RoH profiles determined relatedness among clones. This genome-wide analysis allows us to estimate the number of clones present within each sample and to quantify their relative contributions. The combined analysis of NRAF and RoH allows us to infer whether observed parasite lineages result from a co-transmission or from distinct infection events (superinfection).

Co-transmitted infections, resulting from simultaneous inoculation of genetically related clones following recombination in the mosquito, are characterized by fragmented RoH patterns, with alternating stretches of homozygosity and heterozygosity. In contrast, superinfections, resulting from independent infections of genetically unrelated clones, display an heterozygous genome with no detectable RoH segments (Nkhoma et al. 2020). Note that, within this dataset, the relatively high parasite genetic diversity makes the likelihood of being infected twice with the same strain very low.

Based on the RoH analysis, we identified samples suitable for relatedness analysis (see next section), including monoclonal infections and polyclonal infections containing a dominant clone representing at least 60% of the infection (Pearson et al. 2016). Instead of relying on the conventional McCOIL ≥2 cutoff, which is based on discrete categorical estimates, we derived an empirically informed threshold using our data

Genomes were independently annotated for relatedness suitability by two annotators, after which we performed a nonparametric ROC curve analysis to evaluate the sensitivity and specificity of candidate eMOI thresholds details are provided in supplementary methods **(page 1, paragraph 3, Fig S1**). We selected an empirically optimized threshold of 1.85, which achieved 97% sensitivity for identifying samples suitable for relatedness analysis.

### Parasite Relatedness

To accurately assess the parasite genetic similarity between samples, we estimated pairwise mean posterior probabilities of IBD between genomes or barcodes using hmmIBD, a hidden Markov model-based software relying on meiotic recombination events given a recombination rate of *Plasmodium falciparum* of 13.5 kb/cM (Schaffner et al. 2018).

To estimate IBD, the model requires pairs of homozygous sites between two samples that are referred to as “comparable sites” in this manuscript. These comparable sites are used by hmmIBD to estimate the probability of two samples to be in IBD, which is equivalent to the expected shared fraction of their genomes. With an IBD of more than 0.9, two samples are considered highly related, hence describing the same parasite genotype.

In highly polyclonal samples, the homozygous sites are often dominated by alleles that are nearly fixed in the population, which can artificially increase IBD estimates. To limit this bias, we required a minimum number of comparable sites where at least one sample carries the minor allele; we refer to these as “informative sites”.

Pairwise IBD relationships were visualized as networks using Gephi version 0.10.1 (Bastian et al. 2009). Samples were clustered using the Fruchterman-Reingold layout algorithm, where nodes represent samples and edges represent IBD values above the 0.9 threshold.

### Statistical analyses

All statistical analyses were performed using R software version 4.5.0. Analyses were stratified by site and infection status.

Comparisons of continuous variables (eMOI, FWs, RoH and IBD) across infection status groups (asymptomatic vs. clinical) and age groups were conducted using the Kruskal-Wallis test, with effect sizes estimated using eta-squared based on the H statistic (η^2^[H]) and 95% confidence intervals (Serdar et al. 2021).

Associations between categorical variables were assessed using chi-square tests; specifically, to compare the distribution of drug resistance markers across age groups and by infection status, with post-hoc power analyses to estimate the minimum detectable difference (MDD) between groups.(Mair et al. 2020).

At the genome level, comparison of the distribution of monoclonal, co-transmission, and superinfection classifications across age groups and infection types was performed using Fisher’s exact test, with effect sizes estimated using Cohen’s w (Serdar et al. 2021).

Genetic relatedness networks were visualized in Gephi (version 0.10.1), with barcode clusters arranged using the Fruchterman–Reingold force-directed layout algorithm.

## Results

### Participants and genotyped samples

Between April 2021 and May 2022, 1133 *P. falciparum* confirmed infections were identified from 869 individuals aged between 1 and 91 years old across all 6 villages, as detailed in previous studies (Legendre et al. 2025; Kazanga et al. 2026). Of these, 441 were asymptomatic carriers actively detected during scheduled visits and 692 were clinical malaria episodes. After genotyping and screening for result quality, we obtained 576 high-quality barcodes in Kedougou corresponding to 67 asymptomatic cohort participants, 114 cohort clinical cases, and 394 non-cohort clinical cases; and 153 high-quality barcodes in Kati, corresponding to 92 cohort asymptomatic and 61 cohort clinical case samples **(Fig. 1, Fig. 2)**. ‘Non-cohort clinical cases’ are village inhabitants who were not part of the cohort.

**Fig 1.**
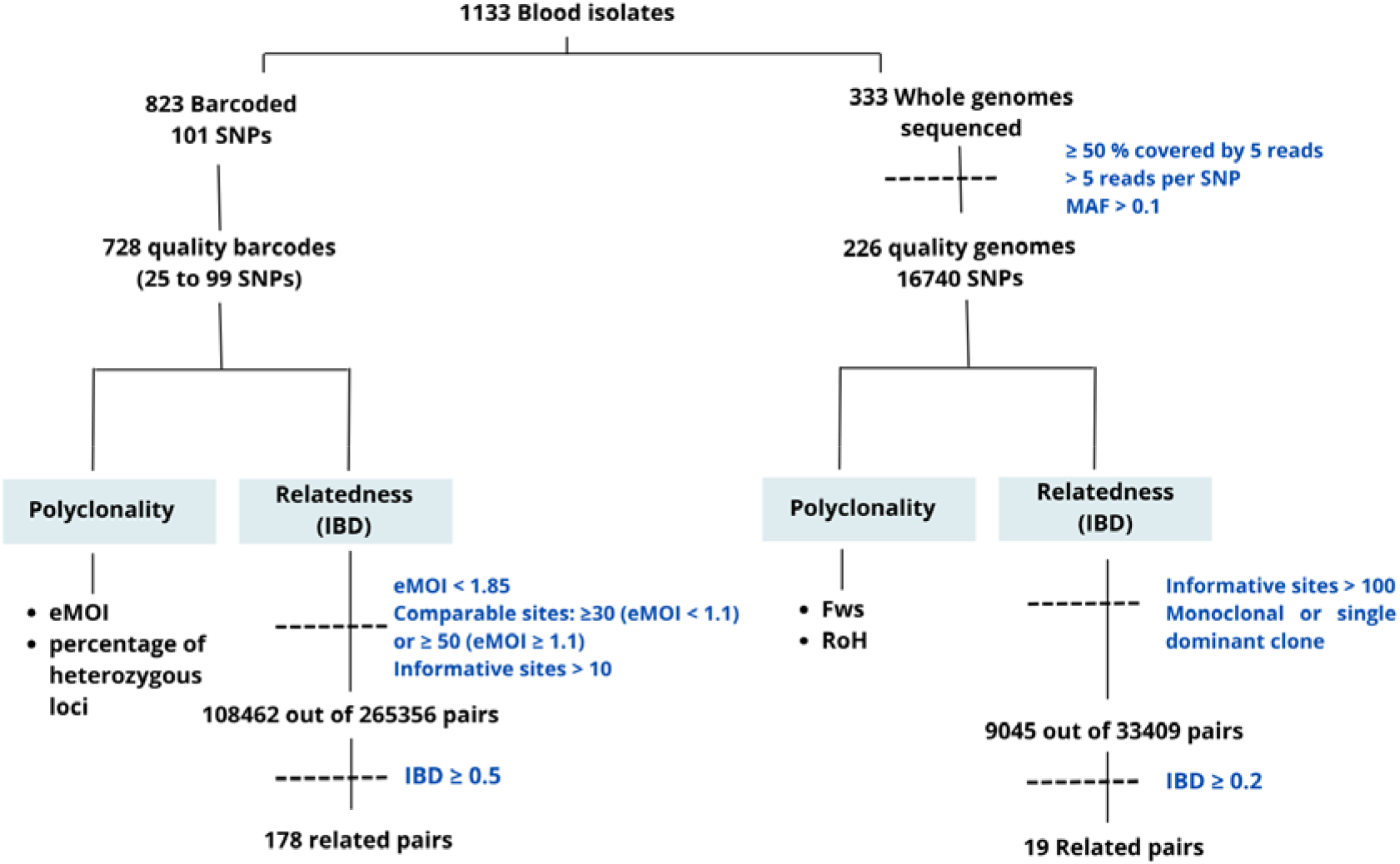
Steps of the combined barcode- and genome-analysis pipeline. Polyclonality was estimated using effective multiplicity of infection (eMOI) and percentage of heterozygous loci on barcodes, and within-host diversity (Fws) and Runs of Homozygosity (RoH) on whole genomes. Relatedness analysis was performed via Identity-by-Descent (IBD) calculation. Genomes were filtered by read depth and Minor Allele Frequency (MAF).

**Fig 2.**
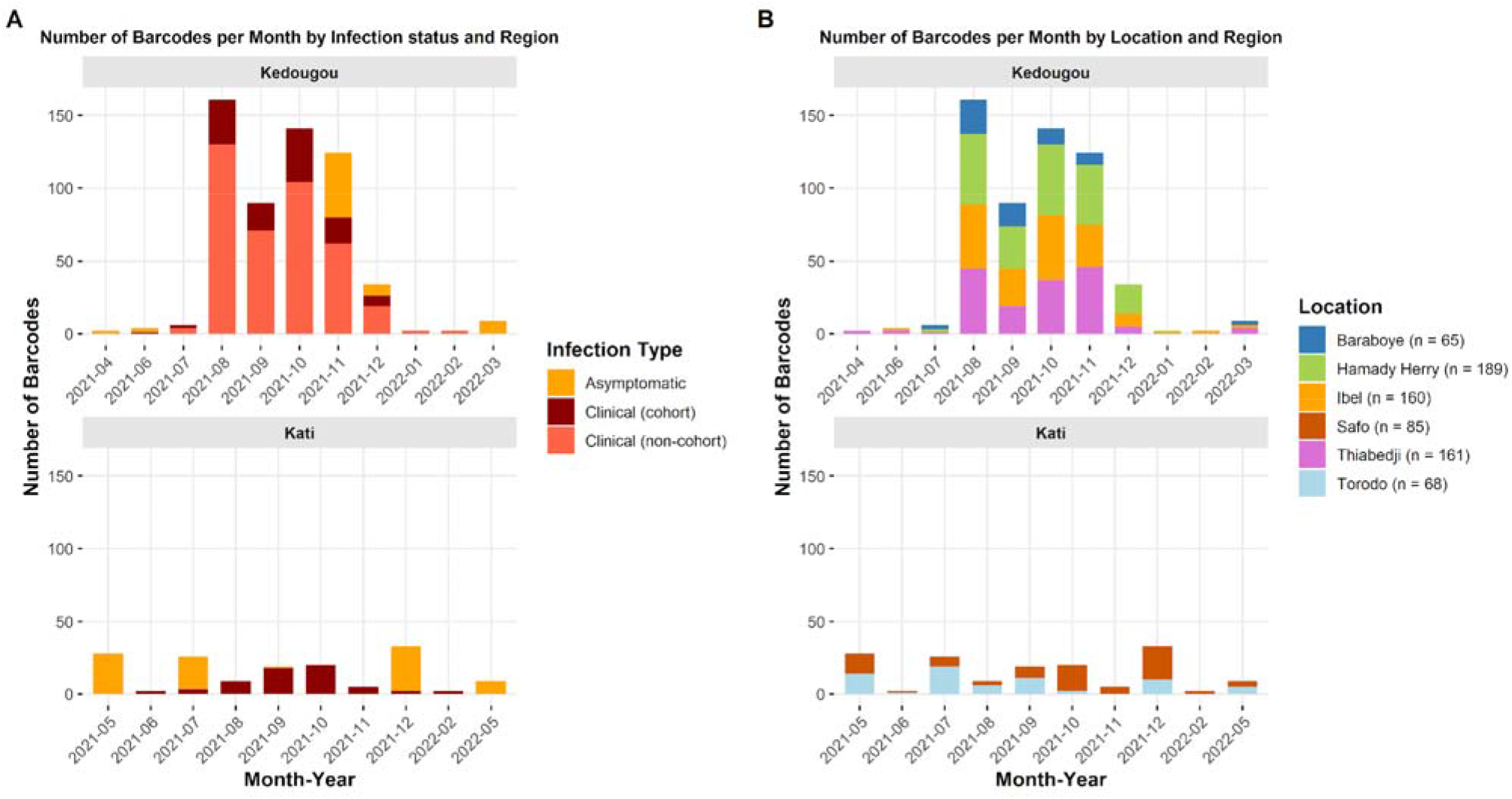
Distribution of high-quality barcodes over time by Infection status (A) and by village (B) in Kedougou (southeastern Senegal) and Kati (southern Mali). Asymptomatic infections were detected during scheduled cohort visits. Clinical infections were identified in cohort participants and non-cohort participants from the same villages (in Kedougou only).

*P. falciparum* genome sequencing on 333 infections resulted in high-quality whole parasite genomes sequences for 200 samples in Kedougou and 26 samples in Kati.

### Barcodes reliably measure MOI

Comparison of molecular and genomic barcodes revealed high concordance **(Fig. S2)**. Agreement rates ranged from 0.82 to 1.00 (median: 0.98, mean: 0.97), confirming the strong reliability of the genotyped barcodes. We measured MOI in genomes with Fws and RoH, and in barcodes with eMOI and proportion of heterozygous loci **(Fig. S3)**. Fws values showed a strong positive correlation with WGS-based RoH (r = 0.90, p < 10^−1^ ⊐) and negative with both barcode metrics: proportion of heterozygous loci (r = -0.96, p < 10^−1^ ⊐) and eMOI (r = -0.94, p < 10^−1^ ⊐).

### Epidemiological patterns of multiplicity of infection

We compared barcode eMOI patterns across infection status (asymptomatic vs. clinical) in both Kedougou and Kati **(Fig. 3)**. Polyclonal infections were defined as those with eMOI > 1.1. In Kedougou, the proportion of polyclonal infections was comparable between asymptomatic (62%, 41/66), clinical cohort (63%, 72/115), and clinical non-cohort (62%, 247/395) cases, with similar median eMOI values (1.19, 1.36, and 1.28, respectively, p = 0.28, η^2^(H) = 0.001, 95% CI [-0.003; 0.02] Kruskal-Wallis test). In Kati, polyclonal infections were more prevalent in asymptomatic individuals (72%, 66/92) compared to clinical cases (56%, 34/61, p = 0.04), although median eMOI did not differ significantly (1.37 vs. 1.14, respectively, p = 0.40, η^2^(H) -0.002, 95% CI [-0.007; 0.04]). eMOI showed no significant association with age groups **(Fig. 4)** in either Kedougou (p = 0.63, η^2^(H) = -0.002, 95% CI [-0.025; 0.005]) or Kati (p = 0.79, η^2^(H) = -0.01, 95% CI [-0.02; 0.08]). These findings suggest that although the proportion of polyclonal infections tends to be higher in asymptomatic carriers in Kati, eMOI values do not differ significantly by clinical status or age group in either setting. The negligible effect sizes suggest that the lack of significance reflects a true absence of meaningful biological difference rather than insufficient statistical power.

**Fig 3.**
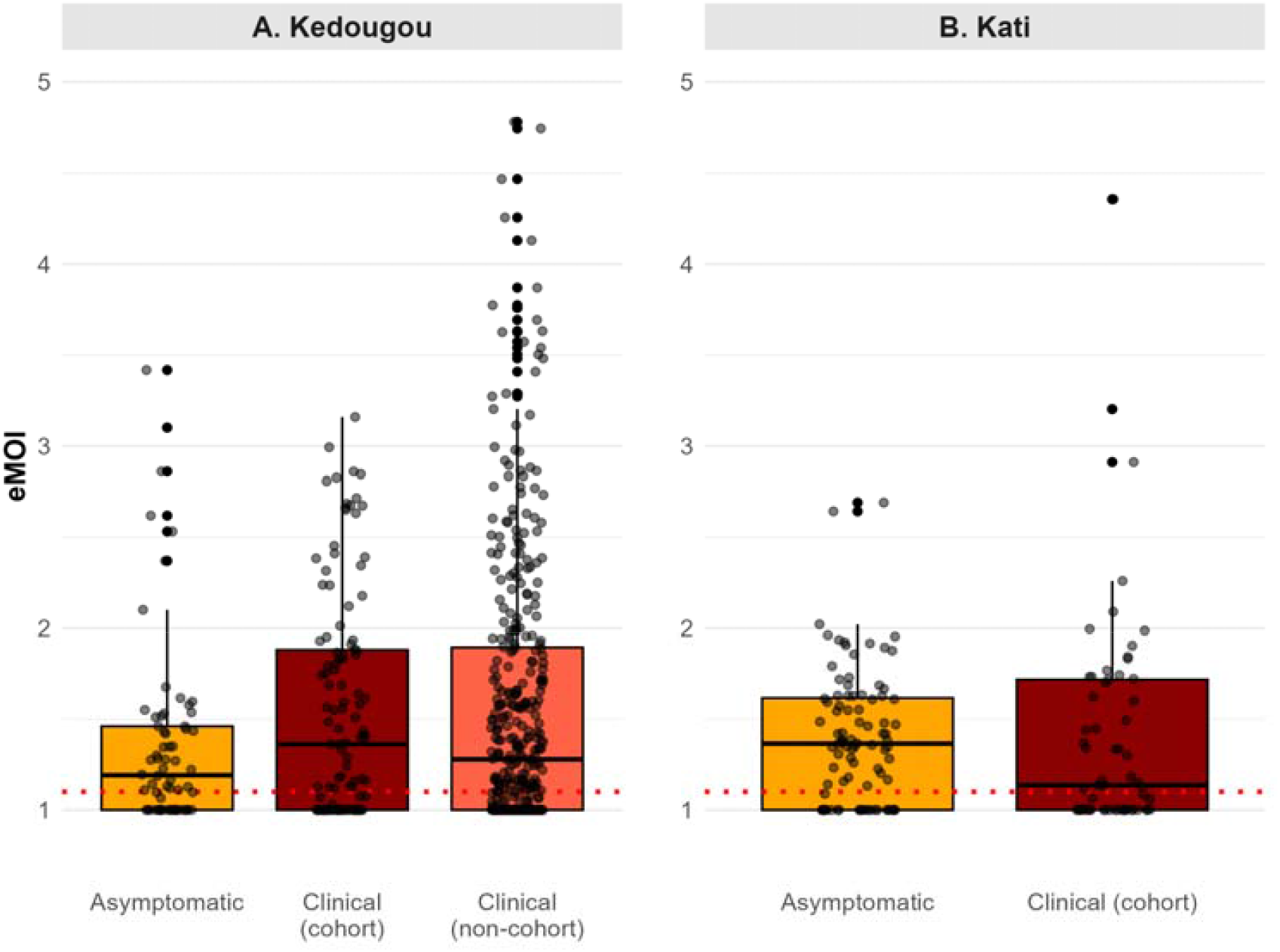
Effective Multiplicity of Infection (eMOI) by Infection status (Clinical vs. Asymptomatic) and by region.

**Fig 4.**
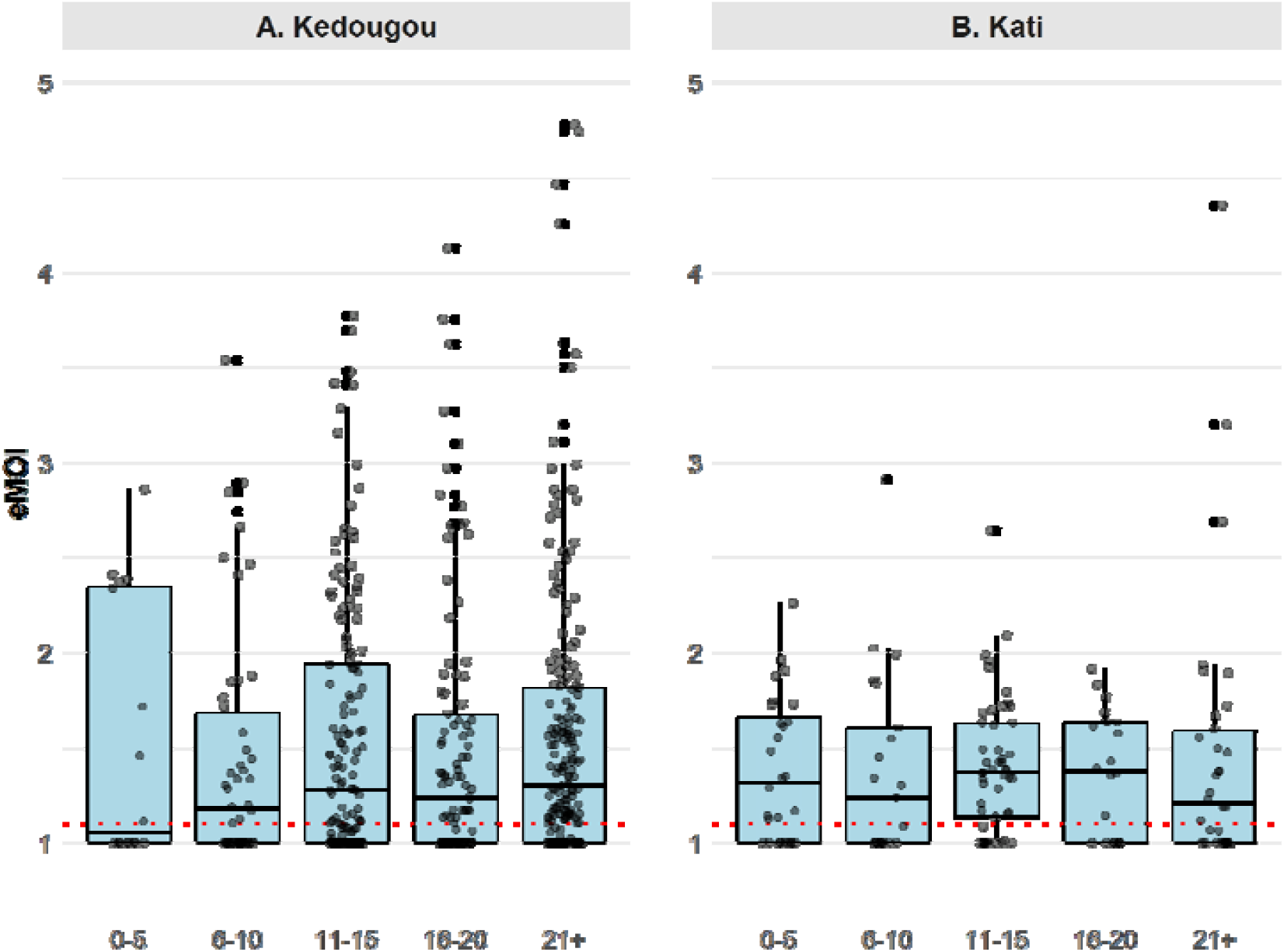
eMOI distribution by age group and by region.

### Characterizing polyclonal infections

RoH patterns revealed distinct infection architectures ranging from monoclonal infections (extensive genome-wide homozygosity) to polyclonal infections with varying numbers and proportions of related or unrelated clones (**Fig S4**.). Of the 226 samples with WGS data, RoH analysis classified 101 (45%) as monoclonal and 124 (55%) as polyclonal. Comparisons between asymptomatic and clinical infections revealed no significant differences in RoH distributions in Kedougou (p = 0.9, η^2^(H) = -0.009, 95% CI [-0.01; 0.03]) or Kati (p = 0.4, η^2^(H) = -0.008, 95% CI [-0.04; 0.25]), despite variation among individual infections within each group **(Fig S5)**.

Among polyclonal infections, 48 (39%) were classified as co-transmission events, while 76 (61%) were classified as superinfections. The distribution across all three polyclonality classes did not differ significantly by infection type in either Kedougou (p = 0.9) or Kati (p = 0.3), nor by age group in either site (p = 0.8 p = 0.2) **(Fig. S6)**. Power analyses indicated power to detect small-to-medium effects in Kedougou (Cohen’s w = 0.23 - 0.24), suggesting no detectable association, while Kati remained underpowered (Cohen’s w = 0.61 - 0.62) and should be interpreted with caution **(Table S1)**.

### Parasite Relatedness

#### Comparison of Barcodes-IBD vs Genomes-IBD

Identity by descent (IBD) was calculated pairwise between 729 high-quality barcodes meeting filtering criteria, yielding 108,462 of 265,356 possible pairs. For the 226 genomes, IBD was estimated for 25,198 out of 25,200 possible pairs. Comparison between barcode-derived and genome-derived IBD was performed on 14,478 pairs with both data types available, identifying a threshold of IBD > 0.5 to define related samples using barcodes **(Fig. S7)**.

#### Parasite from different infection groups can be genetically related

In Kedougou, among 71,784 pairwise barcode comparisons, only 145 (0.20%) demonstrated relatedness with an IBD above 0.5, indicating a highly diverse parasite population. Of these, 86 (59%) were identical (IBD ≥ 0.9), while 59 (41%) were highly related (0.5 ≤ IBD < 0.9) **(Fig. 5A)**. Highly related pairs were significantly more frequent within villages (0.59%) than between villages (0.054%) (p < 0.001), consistent with localized transmission.

**Fig 5.**
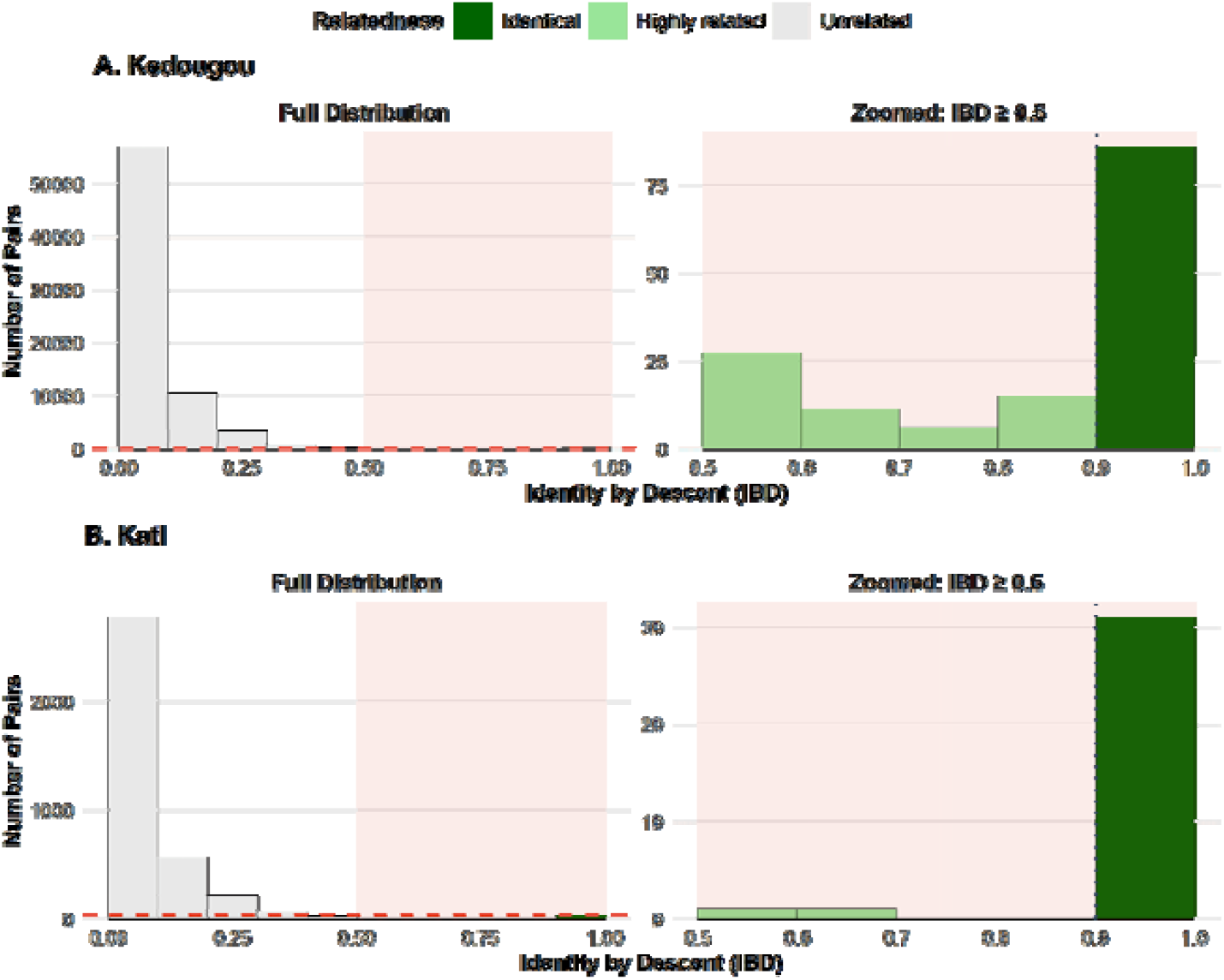
Genetic relatedness in Kedougou (A) and Kati (B). Left panels show the full distribution of barcode pairs, while right panels zoom in on highly related pairs (IBD ≥ 0.5).

A similar pattern was observed in Kati. Among 3,678 pairwise comparisons, 33 (0.90%) showed relatedness (IBD ≥ 0.5), of which 31 (94%) were identical and 2 (6%) were highly related **(Fig. 5B)**. Highly related pairs were also more frequent within villages (1.6%) than between villages (0.01%) (p < 0.001).

Network analysis of highly related pairs revealed transmission clusters comprising both clinical and asymptomatic infections **(Fig. 6)**. No clusters were restricted to a single infection type, indicating that genetically related parasites circulate across infection groups.

**Fig 6.**
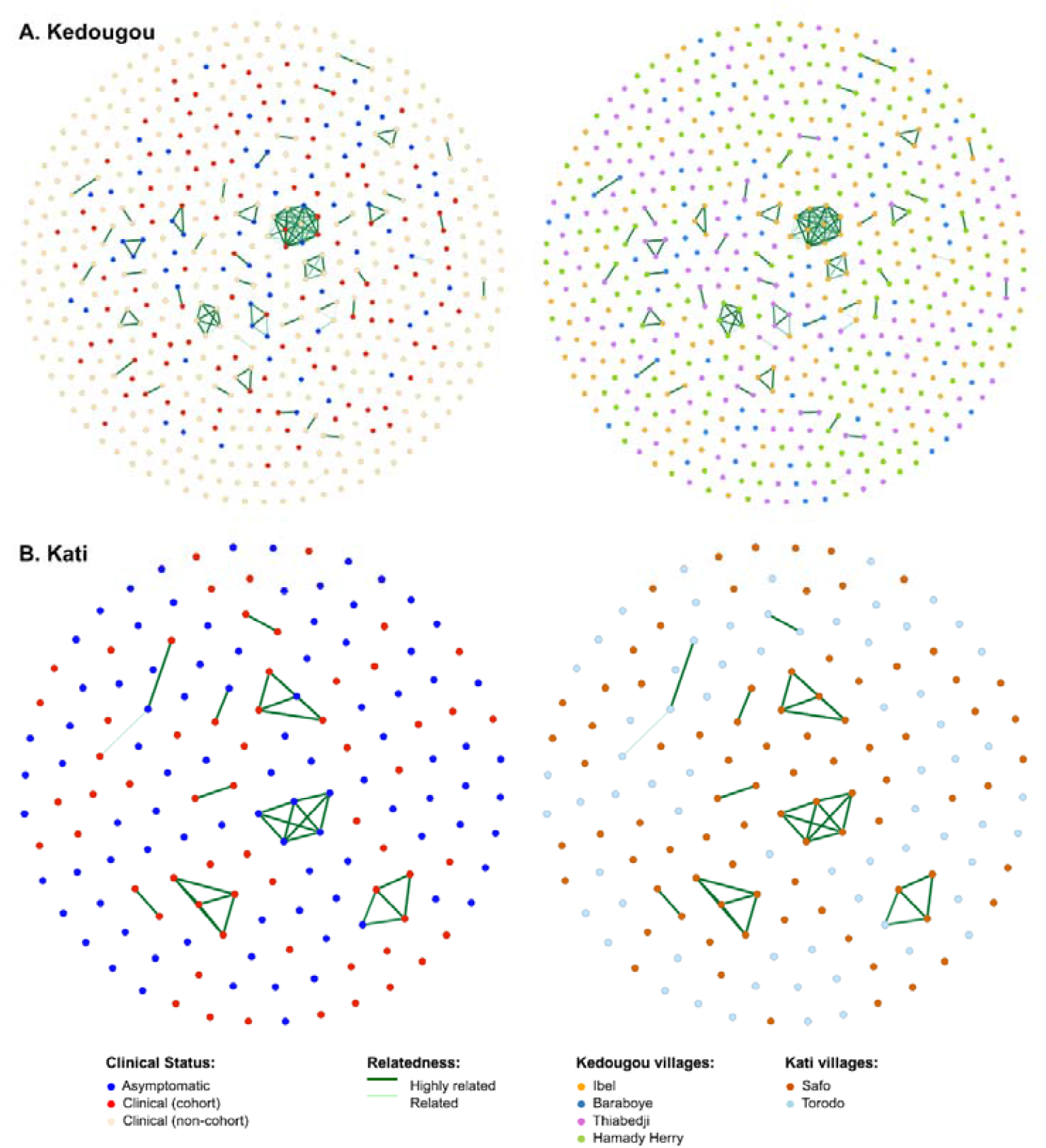
Barcode-IBD networks in Kedougou (A) and Kati (B) colored by infection type (left panels) and by location (right panels). Barcodes are grouped into clusters using the Fruchterman-Reingold layout algorithm in Gephi (version 0.10.1). Networks represent combined cohort and non-cohort samples; cohort-only networks for Kedougou are shown in **(Fig. S9)**.

#### Parasite relatedness decreases with time

In terms of temporality, pairwise relatedness of barcodes declined significantly with time between sampling in both regions. In Kedougou, 0.27% (127/46,589) of pairs sampled within 0–2 months were highly related (IBD ≥ 0.5), compared to 0.07% (18/25,195) of pairs sampled ≥ 2 months apart (p < 0.001). A similar pattern was observed in Kati, where 1.45% (23/1589) of pairs sampled within 0–2 months were related, compared to 0.48% (10/2,089) of pairs sampled ≥ 2 months apart (p = 0.004). These patterns are consistent with ongoing local transmission over short timeframes **(Fig. S8)**.

### Drug resistance

We compared resistance between clinical and asymptomatic infections in both sites by analysing *pfdhfr* and *pfdhps*, key markers of sulfadoxine-pyrimethamine (SP) resistance used for seasonal chemoprevention in these regions (Nwakanma et al. 2014) **(Fig S10 and S11)**. High levels of resistance were observed at both sites: The *pfdhfr* S108N mutation showed very high prevalence, with 96% of isolates carrying this marker in both Kedougou (95% Wilson CI: 0.94-0.97) and Kati (95% Wilson CI: 0.91-0.98). For *pfdhps*, the A437G mutation was present in 80% of isolates in Kedougou (95% CI: 0.76-0.83) and 74% in Kati (95% CI: 0.65-0.81).

No significant association was detected between resistance markers and infection status, except for *pfdhfr* in Kati, where 6.9% of asymptomatic infections carried the wild-type allele compared to none in the clinical group (Fisher’s exact test, p = 0.003). Despite SP administration in children, neither marker showed significant age-related differences in either site. However, the absence of significance should be interpreted with caution, as minimum detectable difference (MDD) analysis revealed that observed differences in resistance prevalence between asymptomatic and clinical infections were small (2.6– 6.4%), yet fell below the MDD (3.3–18.8%), suggesting insufficient statistical power rather than a true absence of biological effect **(Table S2)**.

## Discussion

Across multiple genomic metrics, including multiplicity of infection, runs of homozygosity, identity-by-descent, and drug resistance markers, we found no evidence of detectable genetic differentiation between parasites responsible for clinical disease and those circulating in asymptomatic individuals. These results suggest that clinical malaria cases provide a useful proxy for the broader parasite population sustaining transmission in these settings. We also confirm a population-level relationship between parasite reservoir and clinical cases, centred on, but extending beyond, households.

The high prevalence of polyclonal infections observed in this study further reflects ongoing transmission intensity, as repeated infectious bites increase the likelihood of superinfection with genetically distinct clones (Tessema et al. 2019). The substantial contribution of co-transmission events observed here aligns with recent genomic studies showing that mosquitoes frequently transmit groups of related parasites generated during sexual recombination (Nkhoma et al. 2020). The similar rate of superinfection in clinical and asymptomatic cases, combined with the relatively high MOI, suggests that partially immune individuals may accumulate one or more *P. falciparum* strains asymptomatically before a novel infection precipitates symptomatic disease.

Amidst this high within-host diversity, eMOI did not differ significantly between clinical and asymptomatic infections or across age groups. The relationship between polyclonality, age, and host immunity nevertheless remains complex and context-dependent, with contradictory findings reported across studies. Supporting this, (Mwesigwa et al. 2024) reported high genetic diversity and comparable MOI between asymptomatic and symptomatic individuals in Uganda. Yet higher MOI has also been observed in symptomatic individuals, often associated with higher parasitaemia rather than clinical status itself (Simpson et al. 2023).

In some settings, MOI has been shown to decrease with age, consistent with the development of immunity and reduced susceptibility to superinfection (Niang et al. 2021). In contrast, other studies have reported higher MOI in older individuals or no association with age at all (Eldh et al. 2020). Our study, within this seasonal malaria setting, is consistent with a complexity of infection that primarily reflects exposure intensity rather than host clinical status.

A striking feature of the parasite populations in both Senegal and Mali was their high genetic diversity. Only a small proportion of parasite pairs were genetically related, consistent with extensive recombination and a large effective parasite population size. Parasite relatedness was significantly higher within villages than between villages, indicating predominantly local transmission. Moreover, genetically related infections were more frequently observed within short sampling intervals, consistent with ongoing transmission chains over relatively short timescales. These findings are consistent with previous genomic epidemiology studies showing that malaria transmission often occurs within localized spatial networks such as households and villages (Guery et al. 2025; Brokhattingen et al. 2024; Fola et al. 2024).

Importantly, transmission clusters identified through IBD networks included both asymptomatic carriers and clinical cases. This pattern suggests that parasites circulating in asymptomatic individuals contribute directly to infections that evolve to clinical disease. The absence of clear genetic separation between these infection types supports the view that clinical outcomes are primarily determined by host factors, such as immunity, age, and parasite density, rather than by genetically distinct parasite populations. Our results contrast with a study in southern Zambia where symptomatic infections were genetically distinct and had lower MOI than asymptomatic cases (Searle et al. 2017), likely reflecting substantially lower transmission intensity.

An important limitation of genetic epidemiology studies such as this one is that IBD estimates can only be calculated for isolates with sufficient informative loci, requiring the exclusion of highly polyclonal infections (Taylor et al. 2024). An important remaining challenge is to incorporate highly polyclonal infections in the network of genetic relatedness of parasites, while the proportion of complex infections tends to increase with malaria transmission intensity. Additionally, asymptomatic infections typically present with lower parasitaemia, which may impair detection of minority variants and limit our ability to fully characterize these infections. More sensitive genotyping tools, such as MAD⊐HatTeR should be considered in future studies (Aranda-Díaz et al. 2025).

Our study revealed no significant difference in the prevalence of drug resistance markers between clinical and asymptomatic infections, suggesting that drug-resistant parasites circulate broadly across the population rather than being concentrated in specific epidemiological groups. Importantly, despite the presence of resistance markers in samples from children under five in Mali and under ten in Senegal, current evidence suggests that SMC remains effective, as incidence in children within that age group remains relatively low (Manga et al. 2023; Kazanga et al. 2026). Our findings have important implications for malaria surveillance. Asymptomatic infections are typically characterized by low parasite densities and are usually detected through active community surveys, making sampling difficult and costly. Our results show that parasites obtained from routine clinical surveillance capture the same underlying parasite population as these circulating in asymptomatic carriers. This suggests that clinical samples can provide a reliable and practical source of material for monitoring parasite genetic diversity, population structure, and drug resistance evolution.

At the same time, our results reinforce the important role of asymptomatic infections in sustaining malaria transmission, particularly because these infections often persist for extended periods and may contribute disproportionately to onward transmission (Soumare et al. 2025; Andolina et al. 2021). Consequently, current malaria control strategies may be insufficient: only LLINs specifically target transmission, while interventions such as SMC and improved access to diagnosis and treatment primarily reduce morbidity and mortality in clinical or vulnerable populations. Our findings highlight the need for broader, age-inclusive interventions to effectively reduce the parasite reservoir and interrupt transmission.

## Conclusion

The key finding of our study is that clinical infections reflect the asymptomatic reservoir in both Kedougou and Kati. This suggests that interventions targeting only children, such as SMC, or focusing on the diagnostic and treatment of clinical cases may not be sufficient to interrupt transmission, as asymptomatic carriers of different age groups contribute substantially to local malaria transmission. Complementary and age-inclusive approaches, including strategies such as mass drug administration, may provide an additional means of reducing transmission.

In this context, genetic metrics derived in this study; such as eMOI, IBD, and resistance markers frequencies, provide a valuable baseline for assessing the impact of future interventions and monitoring changes in parasite population structure over time.

## Data Availability

All data produced in the present study are available upon reasonable request to the authors

## Data availability

The barcode data analysis pipeline can be found at https://github.com/marcguery/malaria-barcodes-genomes-gambia. This publication uses data from the MalariaGEN *Plasmodium falciparum* Community Project as described in: An open dataset of Plasmodium falciparum genome variation in 7,000 worldwide samples. Genomes and barcodes data are available in Supplementary files 1 and 2. Whole genome sequencing data are accessible in the European Nucleotide Archive.

## Acknowledgements

The authors would like to thank the staff of MalariaGEN, Wellcome Sanger Institute Sample Management, Genotyping, Sequencing, and Informatics teams for their contribution.

## Funding

Fondation pour la Recherche Médicale FRM EQU202303016290.

Excellence Initiative of Aix-Marseille University—A*MIDEX, a French Investissements d’Avenir programme : projects MARS (A*Midex International 2018) and COCOTIA (AMX-21-IET-017).

---

**Table S1.** Post-hoc power analysis for polyclonality (monoclonal, co-transmission, superinfection) classification by infection status and age group.

| Region | Comparison | Effect size (Cohen's w) | Conclusion |
| --- | --- | --- | --- |
| Kedougou | Polyclonality vs infection type | 0.24 | True absence of effect |
| Kedougou | Polyclonality vs age | 0.23 | True absence of effect |
| Kati | Polyclonality vs infection type | 0.61 | Underpowered |
| Kati | Polyclonality vs age | 0.62 | Underpowered |

**Table S2.** Statistical power analysis for comparisons of *pfdhfr* and *pfdhps* resistance prevalence between asymptomatic and clinical infections in Kedougou and Kati.

| Marker | Region | Asympto (%) | Clinical (%) | Obs diff (%) | MDD (%) | Conclusion |
| --- | --- | --- | --- | --- | --- | --- |
| *pfdhfr* | Kedougou | 98.0% | 95.4% | 2.6% | 3.3% | Underpowered |
| *pfdhfr* | Kati | 93.1% | 100% | 6.9% | - | Significant<br>p=0.003 |
| *pfdhps* | Kedougou | 84.8% | 79.0% | 5.8% | 15.3% | Underpowered |
| *pfdhps* | Kati | 77.6% | 71.2% | 6.4% | 18.8% | Underpowered |

