## Supplementary figures and images for "Clinical malaria cases reflect the parasite population diversity of the asymptomatic reservoir in West Africa"

### Fig. S1

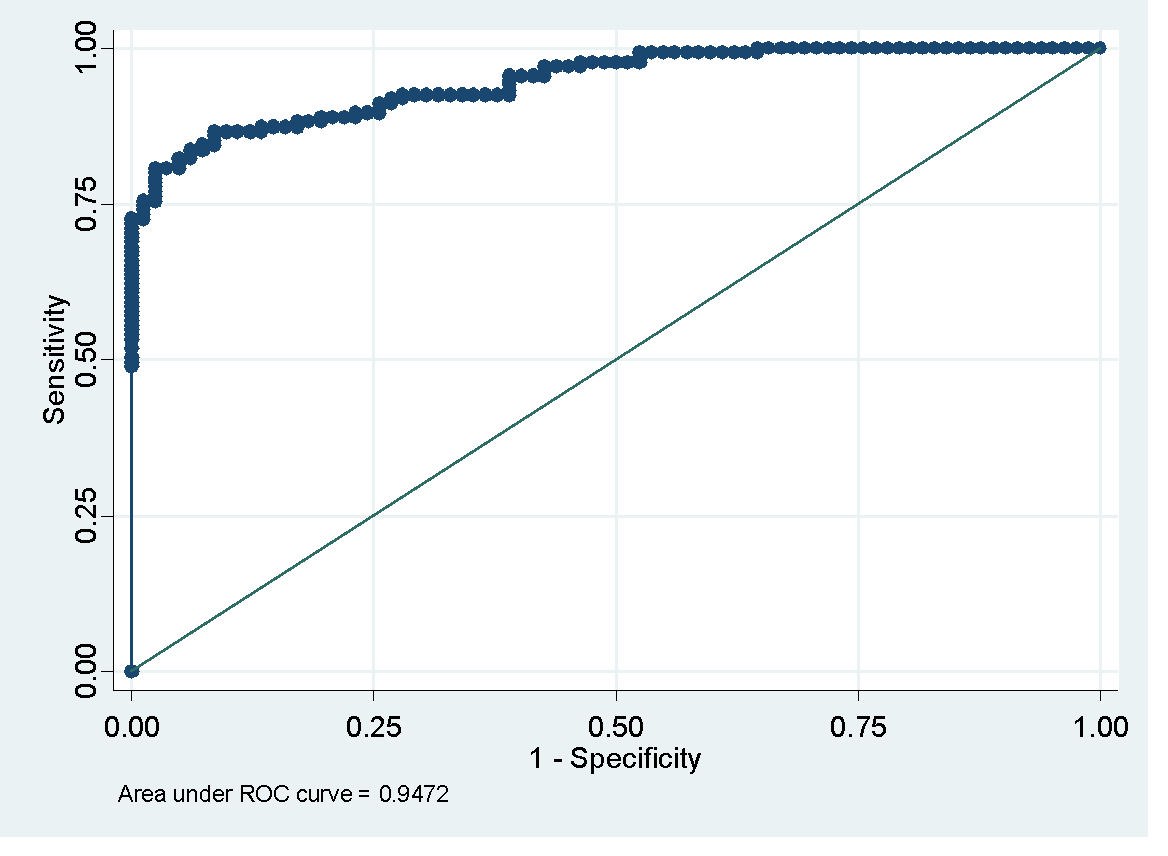

**Fig S1. ROC curve for eMOI threshold selection.** Each dot represents a candidate eMOI threshold.

### Fig. S3

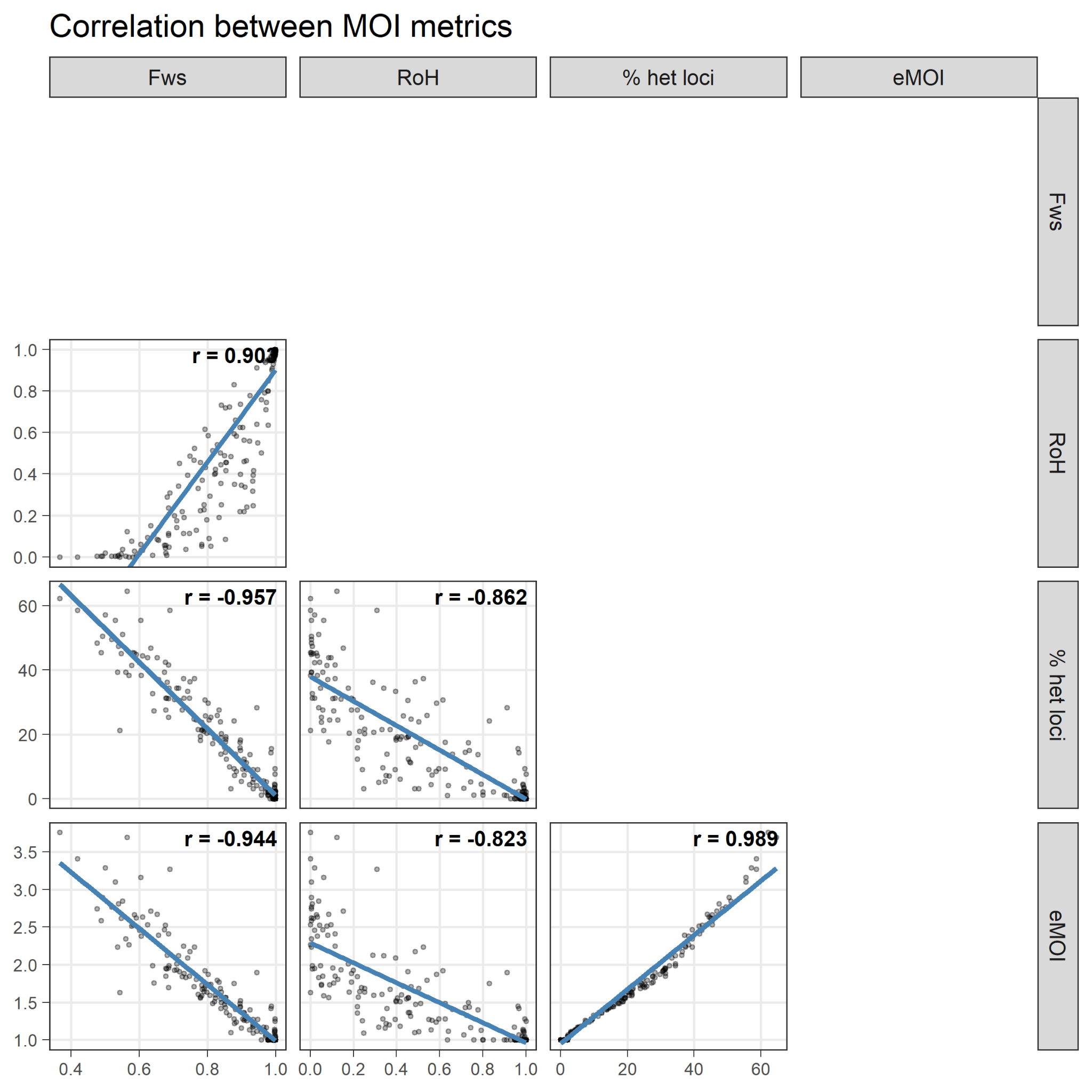

**Fig S3. Correlation between MOI metrics**

### Fig. S5

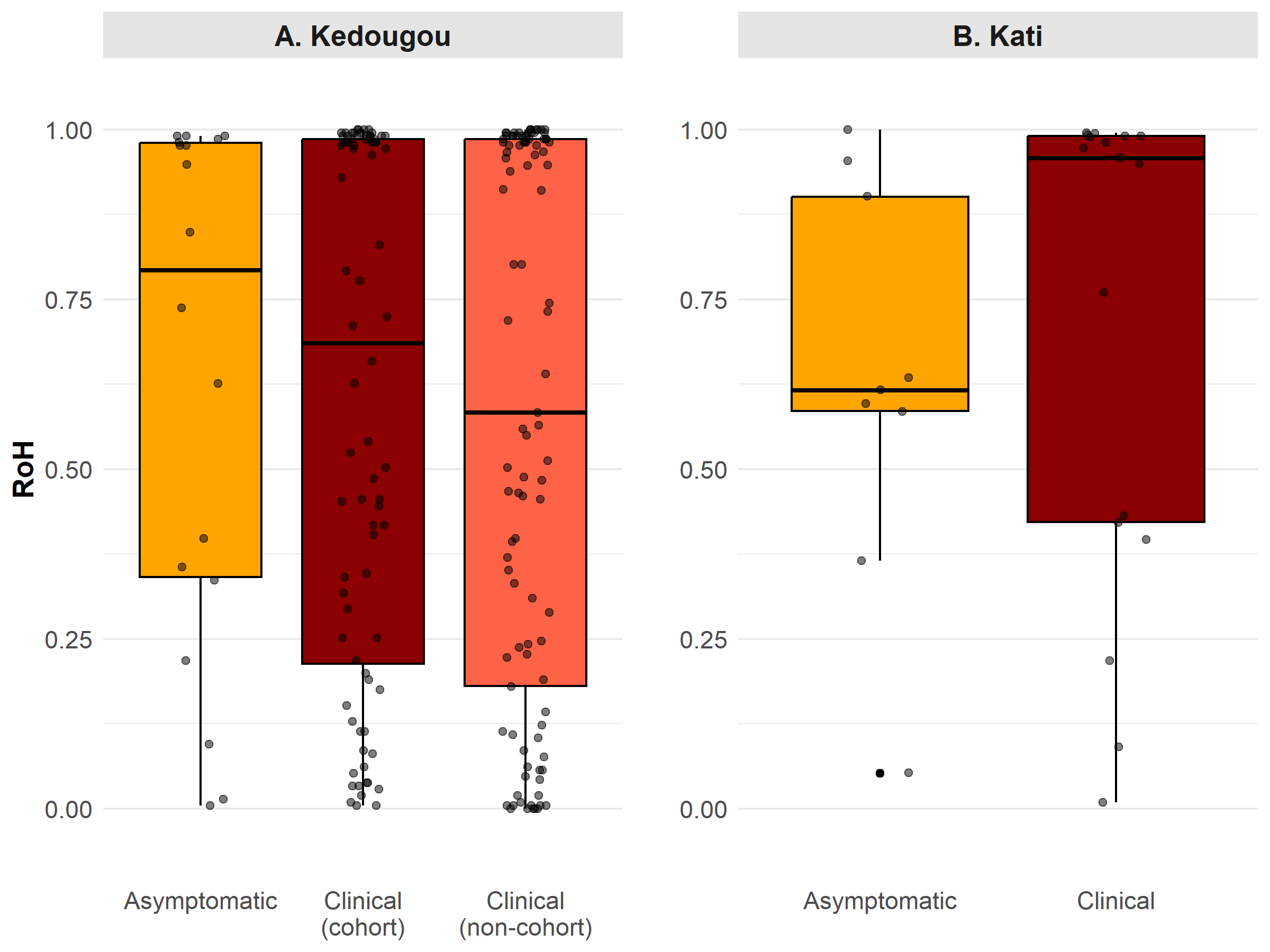

**Fig S5. RoH distribution by Infection status (Clinical vs. Asymptomatic) and by region**
