## Supplementary material for "Clinical malaria cases reflect the parasite population diversity of the asymptomatic reservoir in West Africa": Fig. S2

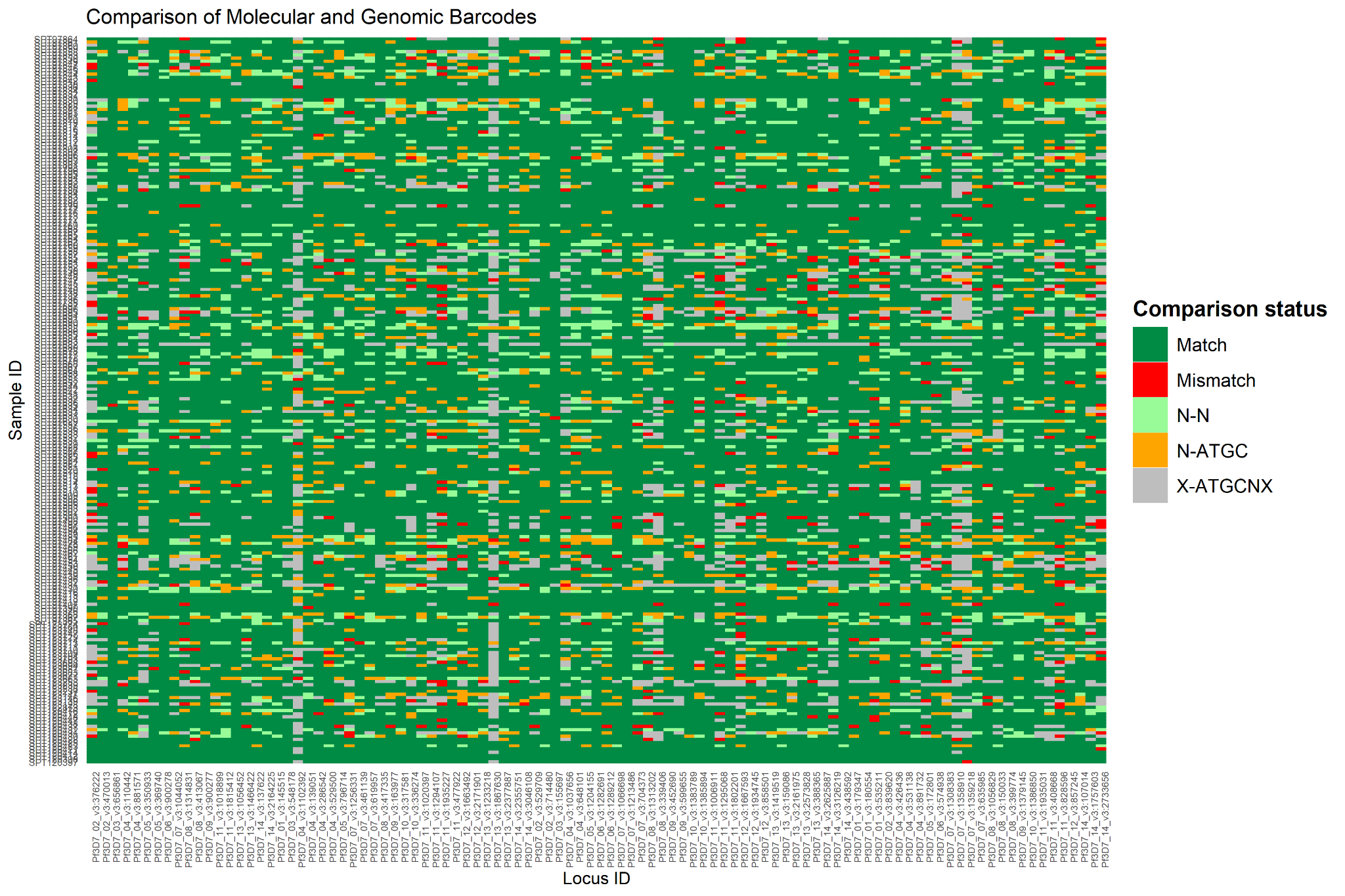

**Fig S2. Comparison matrix of molecular barcodes (constructed from amplicon-based sequencing of 101 SNPs) with genomic barcodes (obtained from whole-genome sequences)**

The agreement rate was calculated as the proportion of matching SNPs among comparable positions:

Agreement = Matches / (Matches + Mismatches). Each row represents a barcode, while each column corresponds to a specific locus.

WGS locus were considered mixed if the within sample MAF was above 0.2.

During quality control, one locus containing only missing values (NAs) in the molecular barcodes and another locus showing a high rate of mismatches with the corresponding genomic barcode were identified and excluded. The remaining 99 high-quality loci were used for subsequent eMOI and IBD analyses.
