## Supplementary material for "Clinical malaria cases reflect the parasite population diversity of the asymptomatic reservoir in West Africa": Fig. S4

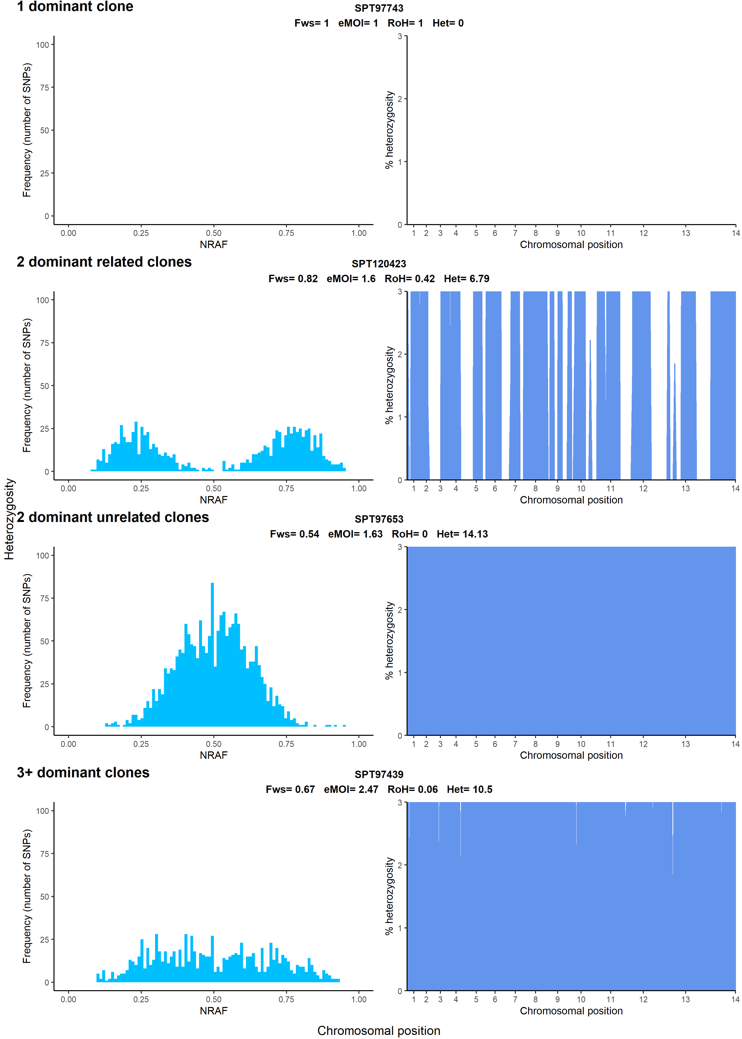

**Fig S4** Illustrative samples showing the non-reference allele frequency (NRAF) distribution across all heterozygous SNPs (left) and heterozygosity (vertical axis) calculated in 20-kb bins with the scale truncated (0–0.03) to highlight RoH. Starting from the top plot, sample 1 (SPT97743) is clonal, as evidenced by Fws and eMOI = 1 and lack of heterozygous SNPs. Samples 2 and 3 contain two dominant clones. Sample 2 contains two related clones, which, based on peaks of NRAFs, are roughly in proportion 80/20. Sample 3 contains 2 unrelated clones (no RoH). Sample 4 appears to contain a complex mixture of unrelated parasites (the relatively flat NRAF distribution indicates multiple dominant clones, with no detectable RoH).
