## Supplementary material for "Clinical malaria cases reflect the parasite population diversity of the asymptomatic reservoir in West Africa": Fig. S6

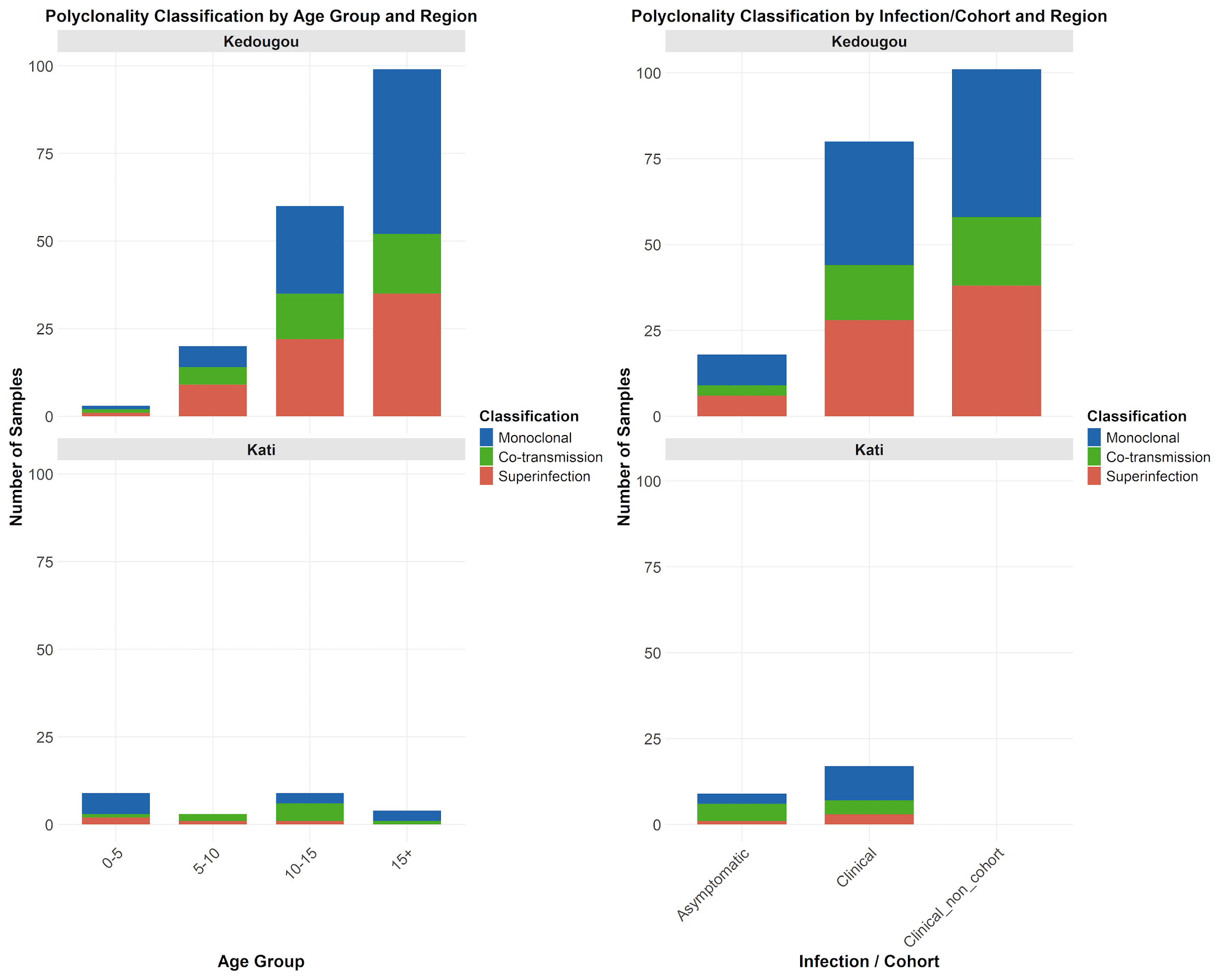

**Fig S6. Polyclonality distribution, based on RoH values, by Infection status (Clinical vs. Asymptomatic) and age by region**
