## Supplementary material for "Clinical malaria cases reflect the parasite population diversity of the asymptomatic reservoir in West Africa": Fig. S7

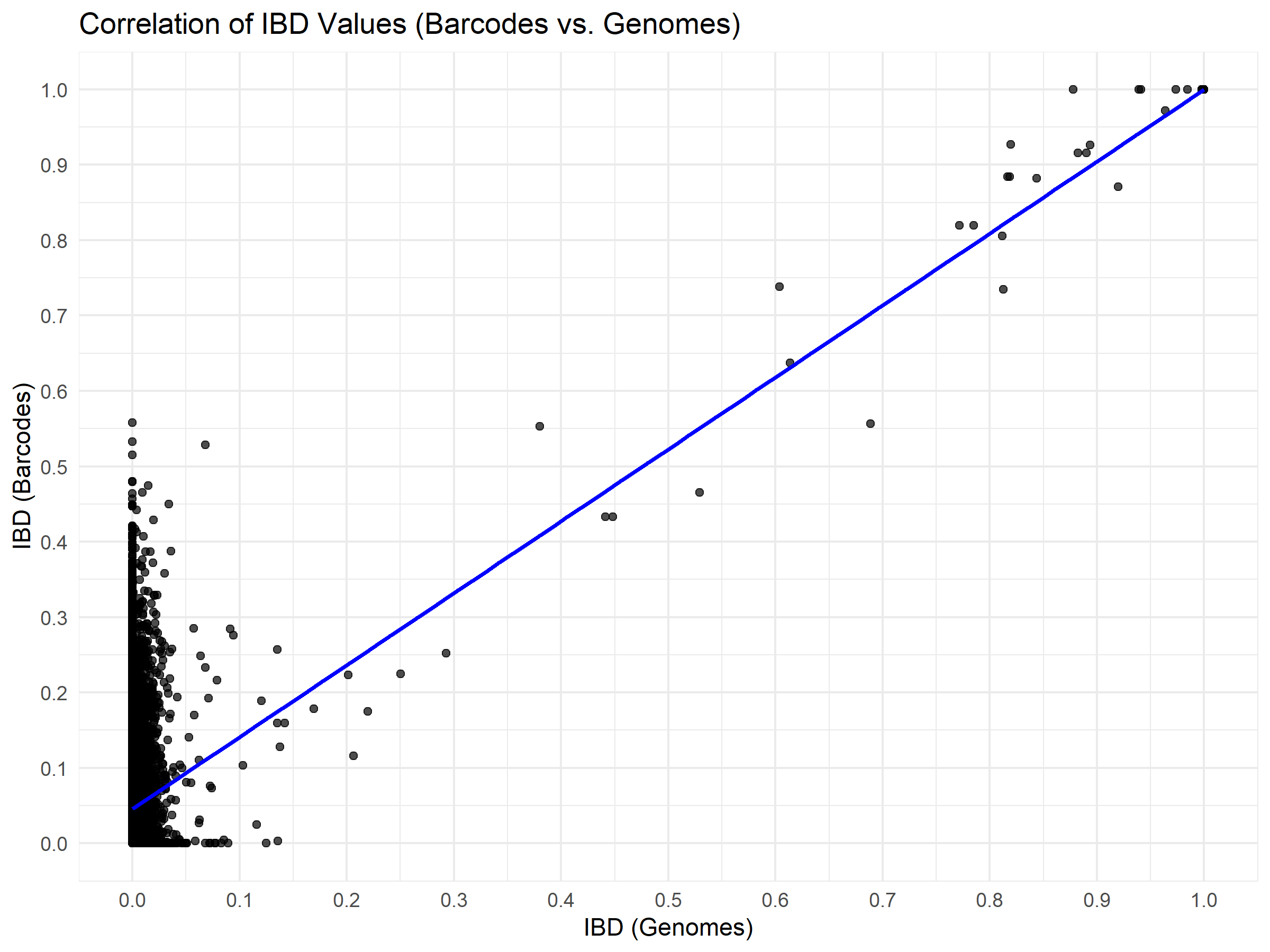

**Fig S7. Comparison of IBD from molecular barcodes and genomic barcodes.** Each dot is a pair of isolates, with their barcode IBD value on the Y-axis and genome IBD value on the X-axis. 226 isolates, with values available for both barcode and genomes, were used for this analysis.

The comparison between barcode-IBD and genome-derived IBD estimates was performed only on the 14478 pairs for which both samples had corresponding barcodes and genomes data. The reliability of barcodes for identifying genetically related isolates was supported by a strong linear correlation between barcode-IBD and genome-IBD, particularly when both were above 0.5 (R^2^ = 0.86, p-value < 10^–15^), with this threshold chosen to distinguish between related (IBD ≥ 0.5) and unrelated (IBD< 0.5) samples for all 576 barcodes. The accuracy of the relatedness classification was then assessed using IBD-Genomes as the gold standard. Overall, the barcode-IBD classification was in strong agreement with the genome-IBD classification (Cohen’s kappa = 0.921) and showed high specificity (1.00), sensitivity (0.97), and precision (0.88).
