## Supplementary material for "Clinical malaria cases reflect the parasite population diversity of the asymptomatic reservoir in West Africa": Fig. S8

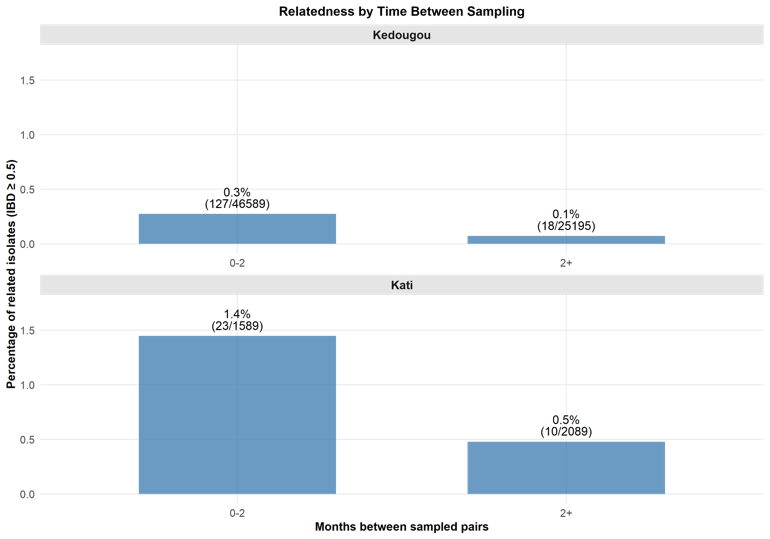

**Fig S8. Percentage of related parasite pairs (IBD ≥ 0.5) by months**

Pairwise relatedness of barcodes decreased significantly with time between sampling in both regions. In Kedougou, 0.27% (127/46589) of pairs sampled within 0–2 months were related (IBD ≥ 0.5, compared to 0.07% (18/25195) of pairs sampled ≥2 months apart (p < 0.001). A similar pattern was observed in Kati, where 1.45% (23/1589) of pairs sampled within 0–2 months were related versus 0.48% (10/2089) of pairs sampled ≥2 months apart (p = 0.004). These results suggest ongoing local transmission within short timeframes
