## Supplementary material for "Clinical malaria cases reflect the parasite population diversity of the asymptomatic reservoir in West Africa": Fig. S9

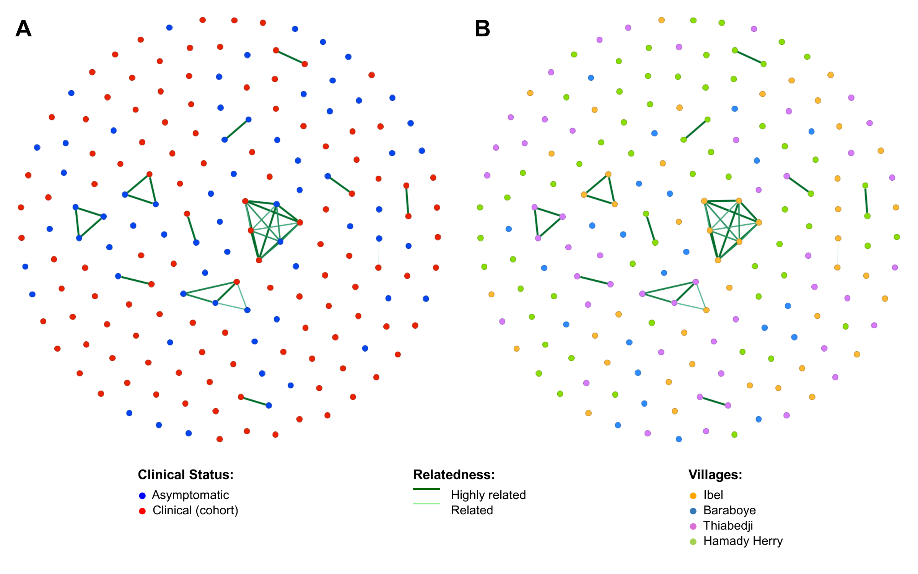

**Fig S9. Kedougou Barcodes-IBD Networks per Infection type (A) and per location (B), cohort samples only**
