## Supplementary material for "Clinical malaria cases reflect the parasite population diversity of the asymptomatic reservoir in West Africa": Fig. S10

| 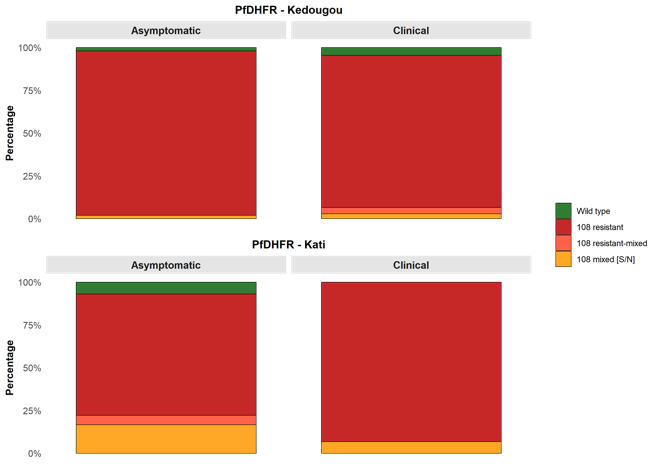 | 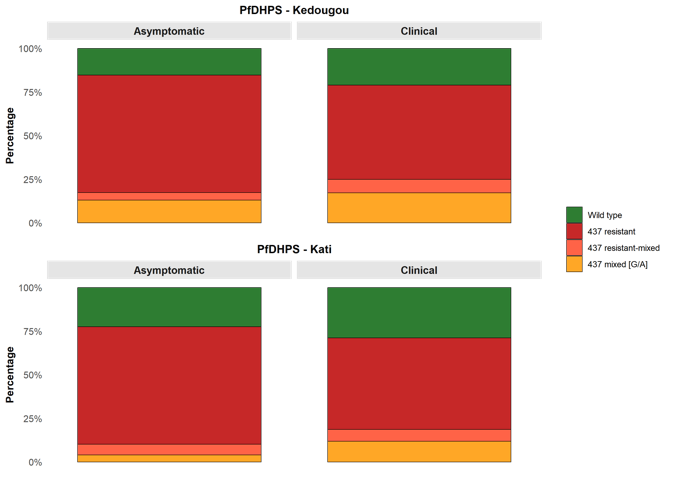 |
| --- | --- |

**Fig.S10 Proportions of *pfdhfr* (left) and *pfdhps* (right) genotypes (Wild-Type, Resistant, Mixed) by clinical status in Kedougou and Kati**

For *pfdhfr*, resistance profiles were not significantly associated with infection status in Kedougou (χ² = 3.007, p = 0.382), but showed a significant association in Kati (χ² = 11.978, p = 0.003). For *pfdhps*, resistance groups showed no significant associations with infection status in either Kedougou (χ² = 3.084, p = 0.382) or Kati (χ² = 3.371, p = 0.324).
