## Supplementary material for "Clinical malaria cases reflect the parasite population diversity of the asymptomatic reservoir in West Africa": Fig. S11

| 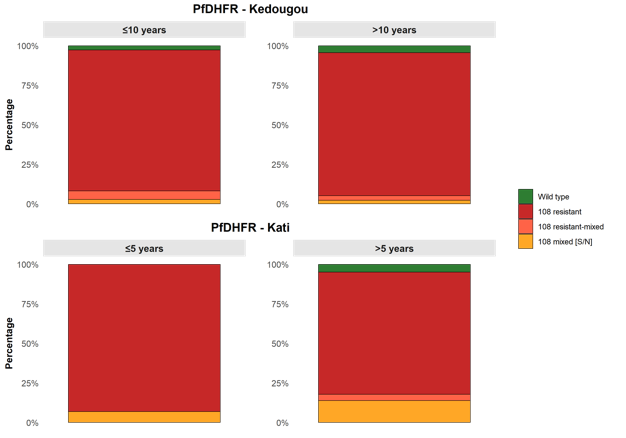 | 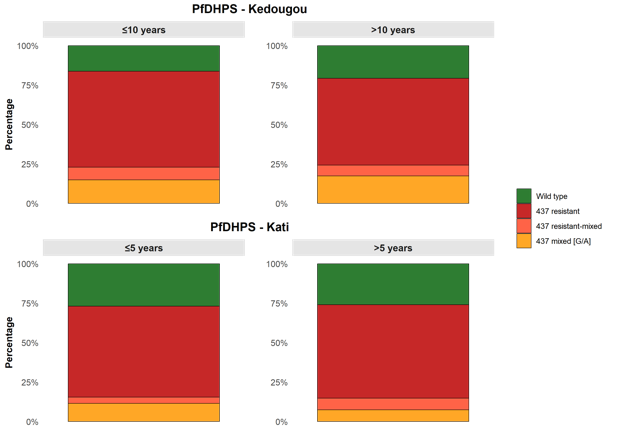 |
| --- | --- |

**Fig. S11 Proportions of *pfdhfr* (left) and *pfdhps* (right) genotypes (Wild-Type, Resistant, Mixed) by age group in Kedougou and Kati.**

Chi-square tests were performed to assess the association between antimalarial drug resistance markers and age groups in each region. For *pfdhps*, no significant associations were observed between resistance groups and age in either Kedougou (≤10 vs >10 years; χ² = 1.3971, p = 0.718) or Kati (≤5 vs >5 years; χ² = 0.7965, p = 0.8545). Similarly, *pfdhfr* resistance profiles showed no significant age-related differences in Kedougou (χ² = 1.797, p = 0.6276) or Kati (χ² = 4.175, p = 0.246).
